# Organ Finder – a new AI-based organ segmentation tool for CT

**DOI:** 10.1101/2022.11.15.22282357

**Authors:** Lars Edenbrandt, Olof Enqvist, Måns Larsson, Johannes Ulén

## Abstract

**Background:** Automated organ segmentation in computed tomography (CT) is a vital component in many artificial intelligence-based tools in medical imaging. This study presents a new organ segmentation tool called Organ Finder 2.0. In contrast to most existing methods, Organ Finder was trained and evaluated on a rich multi-origin dataset with both contrast and non-contrast studies from different vendors and patient populations.

**Approach:** A total of 1,171 CT studies from seven different publicly available CT databases were retrospectively included. Twenty CT studies were used as test set and the remaining 1,151 were used to train a convolutional neural network. Twenty-two different organs were studied. Professional annotators segmented a total of 5,826 organs and segmentation quality was assured manually for each of these organs.

**Results:** Organ Finder showed high agreement with manual segmentations in the test set. The average Dice index over all organs was 0.93 and the same high performance was found for four different subgroups of the test set based on the presence or absence of intravenous and oral contrast.

**Conclusions:** An AI-based tool can be used to accurately segment organs in both contrast and non-contrast CT studies. The results indicate that a large training set and high-quality manual segmentations should be used to handle common variations in the appearance of clinical CT studies.

## 1 Introduction

The application of artificial intelligence (AI) in diagnostic imaging has increased drastically during the last decade. At the centre of this development is a type of models called convolutional neural networks (CNN). We have used CNNs in several projects for automated analysis of positron emission tomography (PET)/computed tomography (CT), and CT studies. The cornerstone in all these projects has been CNN-based organ segmentation in CT.

Organ segmentation can be used to estimate muscle volume, a measure that can be used to predict survival in cancer patients.^1,2^ In PET/CT, CNN-based organ segmentation can be used as a first step in a tumor detection pipeline.^3–5^ A CNN trained to detect tumors takes three 3D images as input, the CT, the PET, and a mask created from the organ segmentation. This organ mask provides anatomical localization for a given PET uptake, making correct classification easier. For example, an uptake well inside the urinary bladder is not likely to be cancer. This approach has been used successfully with, for example, prostate cancer,^3^ lung cancer,^4^ and lymphoma.^5^ One basic organ segmentation tool from RECOMIA, Segmentation tool v1.0, has been used in several research projects.^6^ This tool showed the feasibility of automatically segmenting up to 100 organs in a few minutes. A limitation of the segmentation tool is that only low dose CT without contrast was used for training and testing. This makes the tool perform poorly for organs, such as the kidneys, that look very different in contrast enhanced CT. Hence, the aim of this study was to develop a new CNN for organ segmentation in CT (Organ Finder) using a richer dataset with both non-contrast and contrast studies from different vendors and patient populations. Results will be compared with the performance of Segmentation tool v1.0.^6^

## 2 Methods

### 2.1 Patients

We retrospectively included 1,171 CT studies from seven publicly available CT databases (Table 1). These databases include CT studies from over 10 different hospitals with at least four different scanner manufacturers (GE, Philips, Siemens and Toshiba) and varying fields of view (abdomen, chest, torso, from the top of the skull to mid-thigh). Intravenous contrast in different phases (arterial to late phase) was used in roughly 80% of the studies and oral contrast in 35 %, while circa 15% of the studies had no contrast enhancement.

**Table 1.** Patient material.

| Database | Number of images | Reference |
| --- | --- | --- |
| Task 07 Pancreas | 420 | 7 |
| Task 03 Liver | 52 | 7 |
| ACRIN 6668 | 203 | 8–10 |
| CT Lymph Nodes | 176 | 8, 11–13 |
| C4KC-KiTS | 170 | 8, 14, 15 |
| CT-ORG | 117 | 8, 16–18 |
| Anti-PD-1 Immunotherapy Lung | 33 | 8, 19 |
| **Total** | 1,171 |  |

A test set was selected to include 20 CT studies (10/10 female/male patients) of which 5 were non-contrast studies, 5 intravenous contrast only, 5 oral contrast only, and 5 with both intravenous and oral contrast. The remaining 1,151 CT studies were used to train the CNN.

### 2.2 Manual Segmentations

Professional annotators manually segmented organs using a cloud-based annotation platform. The platform includes basic display features for segmentation of CT studies, for example drawing tools for predefined Hounsfield unit intervals. Every segmentation was first checked by a dedicated quality person and returned to the annotator if adjustments were necessary. A final quality check was done by an experienced nuclear medicine specialist and segmentations with insufficient quality were returned to the annotation team.

Twenty-two different types of organs were segmented, but not all organs were segmented in all CT studies. The left and right kidney, lung, femur, humerus, and hip bone were segmented separately. Left and right clavicles, iliac arteries, and scapulae were combined to one segmentated organ. The ribs and spine were segmented as one organ each. The final database of segmentations consisted of 5,826 quality checked organs (3,856 in intravenous contrast CT and 1,970 in CT without intravenous contrast) (Table 2).

**Table 2.**
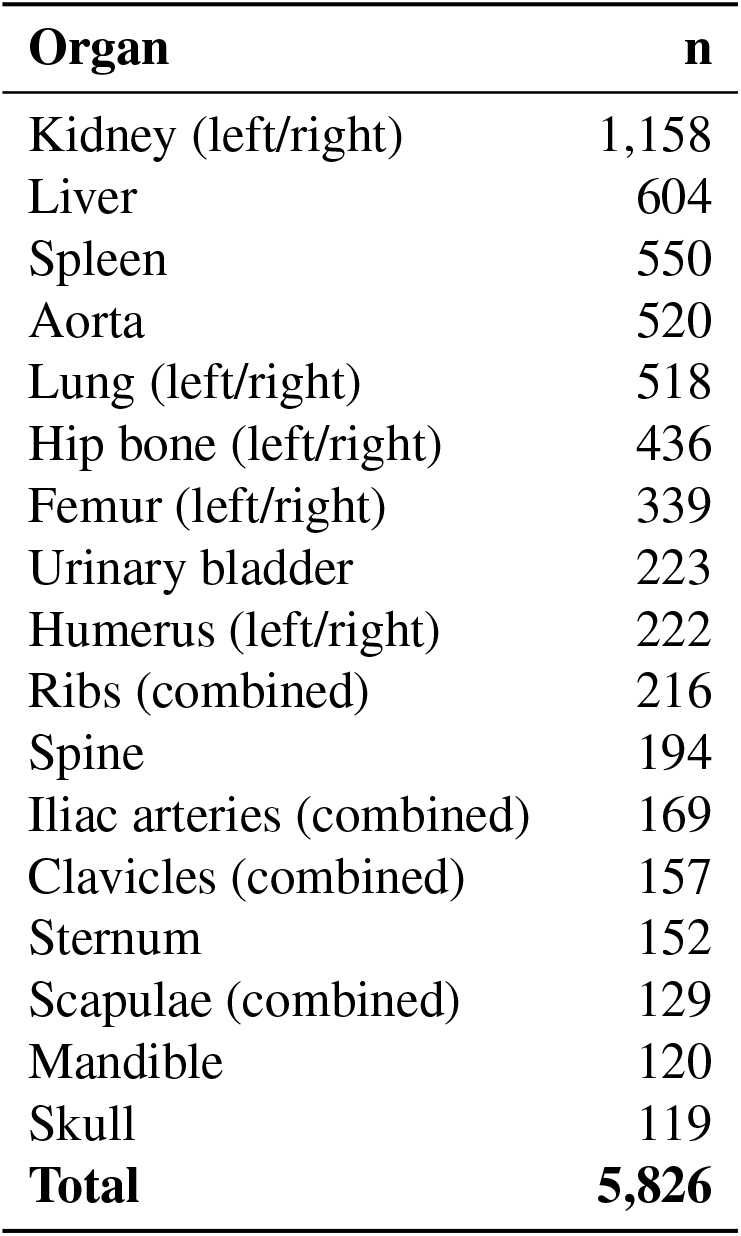
Segmented organs.

## 3 AI tool

### 3.1 Network

The CNN used for organ segmentation is a 3D U-Net.^20^ It uses convolutions, max pooling and transposed convolutions to process the input CT image on four different resolutions. The information, or features, from lower resolutions are upsampled using transposed convolutions and concatenated with higher resolution features via skip connections across the encoder and decoder part of the CNN, see Fig. 1. For an input volume of 100 *×* 100 *×* 100 voxels the CNN outputs an estimated class probability for each voxel of a 12 *×* 12 *×* 12 volume at the center of the input volume. Before input to the CNN, the CT is resampled to a voxel size of 1 *×* 1 *×* 3 mm. The intensities are clamped to [*−*800, 800] and then rescaled to [*−*1, 1].

**Fig. 1.**
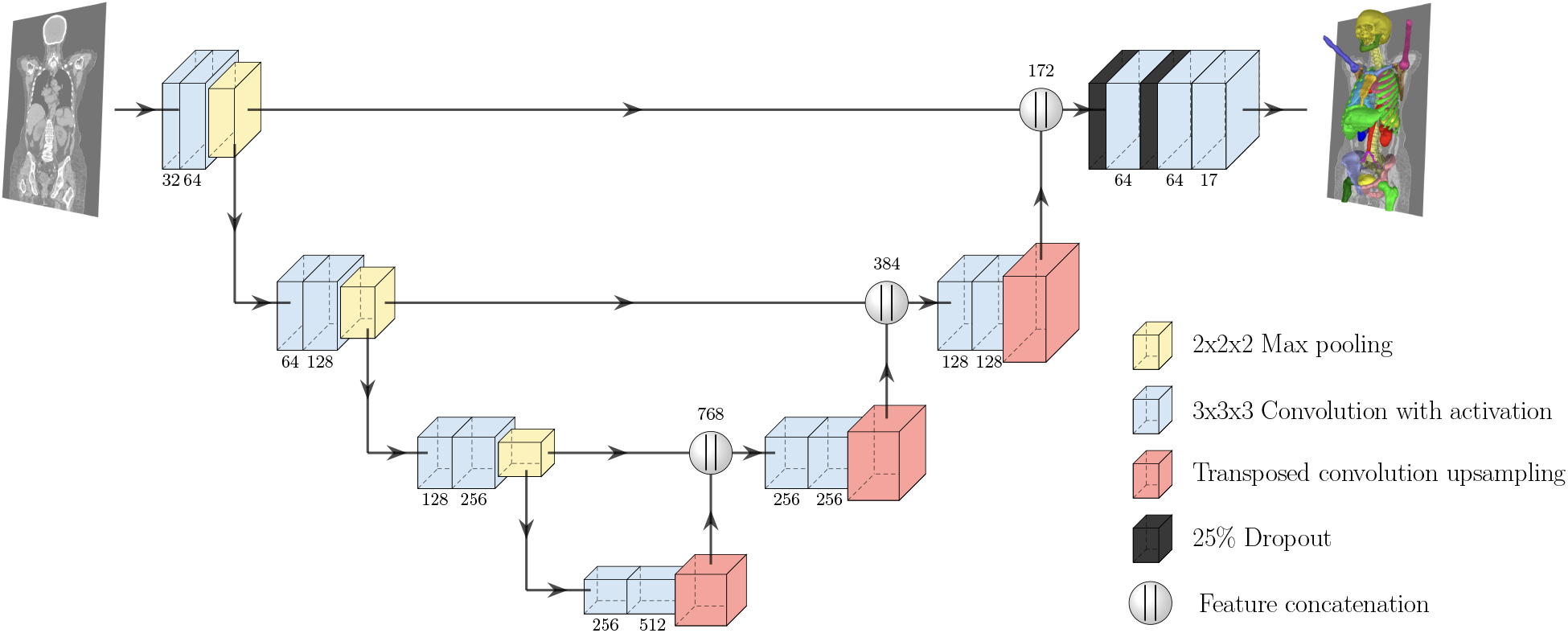
Schematic of the 3D U-Net, each layer or operation is represented by a box and connections are visualized with arrows. When applicable, the number of features maps output are shown below (or above) each layer (or operation). All convolutions are done without padding, hence cropping is needed before feature concatenation. Both downsampling with max pooling and upsampling with transposed convolution scales the spatial dimension with a factor two.

### 3.2 Sampling and Training

Training was done with categorical cross-entropy loss and Adam^21^ optimization. The learning rate followed the exponential decay schedule with initial learning rate 10^*−*4^ and a decay rate of 0.95 every 10^5^ samples. Sampling was focused on foreground voxels and background close to foreground since this generally helps the CNN to learn to segment the edges of organs correctly. The different organs were initially sampled evenly but every 5 *·* 10^5^ training steps, the current model was run on all images in the training set and voxels with high loss was later sampled more frequently.

In addition to applying the cross-entropy loss on the final output layer, deep supervision was used, meaning that the loss was applied on all depth levels of the network. The loss terms were weighted with 1.0, 0.75, 0.5 and 0.25 (highest to lowest scale). For augmentation, a random scaling and rotation was applied to the input and target pair. In addition, Gaussian noise was added to the input images.

### 3.3 Inference

During inference, the CNN, due to being fully convolutional, can be efficiently applied on the entire CT image outputting estimated class probabilities for every voxels. An initial segmentation is then created by taking the argmax for each voxel. Postprocessing is done independently for each organ. For most organs all segmented voxels except the largest connected component is removed and holes in the segmentation are filled. For iliac arteries, clavicles, scapulae and sternum the two largest components are kept and for the ribs, all components larger than 1 ml are kept.

## 4 Statistical Analysis

The AI-based segmentations were compared to the corresponding manual segmentations of the test set. The Sørensen-Dice (Dice) index was used to evaluate the agreement between automated and manual segmentations by analysis of the number of overlapping voxels. For each organ, a (two-sided) sign test was used to test whether there was a significant difference in Dice between Organ Finder and Segmentation tool v1.0.

## 5 Results

The new AI-tool for organ segmentation in CT (Organ Finder 2.0, SliceVault AB, Sweden) was applied to the 20 CT studies of the separate test set. One of the test patients is shown in Figure 2. The CNN-based segmentations were compared to the corresponding manual segmentations. Per organ metrics are shown in Table 3. The results for the left and right kidneys, lungs, femurs, humeri, and hip bones are grouped. The average Dice index over all organs was 0.93. The AI-tool showed high performance for all four subgroups of the test set based on presence or absence of intravenous and oral contrast. The average Dice index was in the interval 0.90 to 0.98 for all bones and all but two soft tissue organs. The most difficult organs to segment manually, the iliac arteries, showed the lowest Dice index of 0.80 for the CT with intravenous contrast. This organ was not possible to manually segment in the non-contrast CT. Unfortunately, the skull and mandible were not present in the contrast CT studies.

**Table 3.** Dice index per organ in the test set (n=20). Each group (iv+oral, iv, oral, non-contrast consisted of 5 patients (iv-intravenous)

| Organ | iv+oral contrast | iv contrast | oral contrast | non-contrast | Total |
| --- | --- | --- | --- | --- | --- |
| Aorta | 0.93 | 0.93 | 0.91 | 0.90 | 0.92 |
| Clavicles | 0.89 | 0.87 | 0.94 | 0.93 | 0.91 |
| Femur | 0.98 | 0.98 | 0.98 | 0.97 | 0.98 |
| Hip bone | 0.97 | 0.97 | 0.97 | 0.96 | 0.97 |
| Humerus | 0.95 | 0.97 | 0.92 | 0.87 | 0.92 |
| Iliac arteries | 0.80 | 0.80 |  |  | 0.80 |
| Kidney | 0.96 | 0.95 | 0.93 | 0.95 | 0.95 |
| Liver | 0.96 | 0.97 | 0.96 | 0.96 | 0.96 |
| Lung | 0.98 | 0.98 | 0.98 | 0.98 | 0.98 |
| Mandible |  |  | 0.89 | 0.91 | 0.90 |
| Ribs | 0.89 | 0.90 | 0.93 | 0.90 | 0.90 |
| Scapulae | 0.94 | 0.94 | 0.95 | 0.94 | 0.94 |
| Skull |  |  | 0.92 | 0.92 | 0.92 |
| Spine | 0.91 | 0.94 | 0.93 | 0.90 | 0.92 |
| Sternum | 0.93 | 0.94 | 0.94 | 0.82 | 0.91 |
| Spleen | 0.95 | 0.98 | 0.95 | 0.96 | 0.96 |
| Urinary bladder | 0.84 | 0.75 | 0.89 | 0.82 | 0.82 |
| **Total** | **0.92** | **0.92** | **0.94** | **0.92** | **0.93** |

**Fig. 2.**
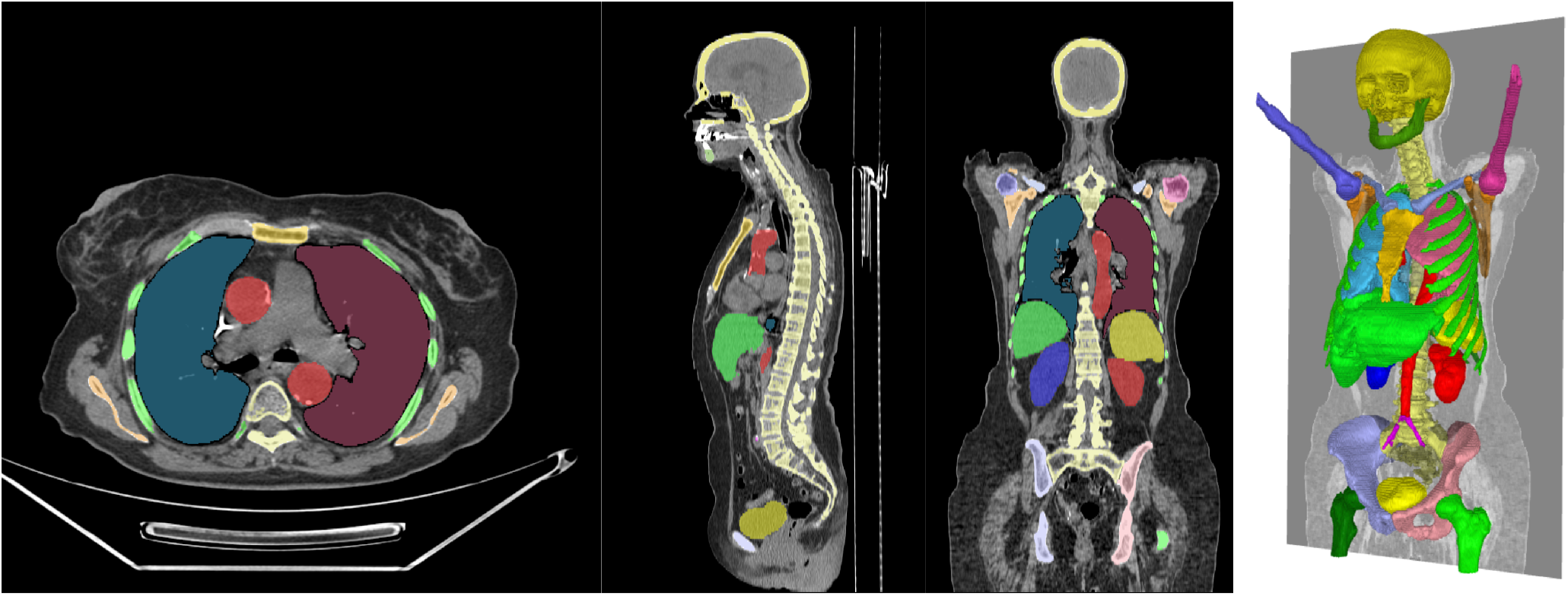
AI-based segmentation of organs in one of the test patients. The average Dice index for all organs was 0.95.

Table 4 shows a comparison between organ segmentation of Organ Finder and Segmentation tool v1.0.^6^ Segmentation tool v1.0 shows much higher Dice index for the test CT studies without intravenous contrast compared to those with contrast (0.90 vs. 0.77). Organ Finder showed significantly (*p <* 0.01) higher Dice index than Segmentation tool v1.0 for 14 of the 15 organs. The only exception was the urinary bladder having higher Dice index in 13 of the 20 patients (*p* = 0.18).

**Table 4.** Comparison Organ Finder vs. Segmentation tool v1.0 in the test set (n=20) (iv-intravenous).

| Organ | with iv contrast |  | without iv contrast |  |
| --- | --- | --- | --- | --- |
|  | Organ Finder | Segmentation tool v1.0 | Organ Finder | Segmentation tool v1.0<sup>6</sup> |
| Aorta | 0.93 | 0.87 | 0.91 | 0.86 |
| Clavicles | 0.88 | 0.49 | 0.94 | 0.91 |
| Femur | 0.98 | 0.96 | 0.97 | 0.97 |
| Hip bone | 0.97 | 0.93 | 0.97 | 0.93 |
| Humerus | 0.96 | 0.34 | 0.89 | 0.89 |
| Kidney | 0.95 | 0.61 | 0.94 | 0.90 |
| Liver | 0.96 | 0.92 | 0.96 | 0.94 |
| Lung | 0.98 | 0.97 | 0.98 | 0.97 |
| Mandible |  |  | 0.90 | 0.84 |
| Ribs | 0.90 | 0.81 | 0.91 | 0.85 |
| Scapulae | 0.94 | 0.84 | 0.95 | 0.89 |
| Skull |  |  | 0.92 | 0.90 |
| Sternum | 0.93 | 0.82 | 0.88 | 0.86 |
| Spleen | 0.96 | 0.79 | 0.96 | 0.95 |
| Urinary bladder | 0.80 | 0.70 | 0.85 | 0.84 |
| **Total** | **0.92** | **0.77** | **0.93** | **0.90** |

## 6 Discussion and Conclusions

Organ Finder was trained to perform organ segmentation in contrast as well as non-contrast CT. The test results show that this aim was accomplished with average Dice index over all organs of 0.93. The importance of including both contrast and non-contrast studies in a training group was illustrated by the results of Segmentation tool v1.0. The Dice index in the subgroup with intravenous contrast was much lower than the index in the subgroup without intravenous contrast (0.77 vs. 0.90).

Organ Finder was significantly better than Segmentation tool v1.0 even in the subgroup without intravenous contrast (0.93 vs. 0.90). One reason for this increased performance is probably the size difference between the training sets (Organ Finder 1,151 vs Segmentation tool v1.0 339). A large training set is needed to sufficiently capture variations in appearance due to, for example, patient gender, age, body size and shape, as well as abnormalities due to different diseases. Technical variation, for example, due to presence or absence of intravenous and oral contrast, and differences in field of view should also be represented in a training set. In addition we included CTs from different vendors as well as patient populations from several hospitals in the United States and Europe in the training set.

The superior performance of Organ Finder is most likely also due to higher quality of the manual segmentations. Written specifications on how different organs should be segmented, professional annotators, and routines to check segmentation quality for every organ by two dedicated persons contributed to a training set of high quality.

A limitation of Organ Finder is that only 22 organs are included compared to 100 for Segmentation tool v1.0. We decided to limit the number of bones as we found that the value of an AI tool separating individual ribs or vertebrae is limited in most clinical applications. Organ Finder segments all bones from the top of the skull to mid-thigh using 13 labels, which can be compared to the 77 different bones included in Segmentation tool v1.0. Future work with Organ Finder will include adding more soft tissue organs for example the gastrointestinal tract, heart, muscle, and prostate.

In conclusion, an AI-based tool can be used to accurately segment organs in both contrast and non-contrast CT studies. The results show that a large training set and high-quality manual segmentations should be used to handle common variations in the appearance of a CT in clinical routine.

## Data Availability

The datasets analysed during the current study are not publicly available due to ethical considerations.

## References

1 P. Borrelli, R. Kaboteh, O. Enqvist, et al., “Artificial intelligence-aided ct segmentation for body composition analysis: a validation study,” European Radiology Experimental 5(1), 1–6 (2021).

2 T. Ying, P. Borrelli, L. Edenbrandt, et al., “Automated artificial intelligence-based analysis of skeletal muscle volume predicts overall survival after cystectomy for urinary bladder cancer,” European Radiology Experimental 5(1), 1–8 (2021).

3 E. Trägårdh, O. Enqvist, J. Ulén, et al., “Freely available, fully automated ai-based analysis of primary tumour and metastases of prostate cancer in whole-body [18f]-psma-1007 pet-ct,” Diagnostics 12(9), 2101 (2022).

4 P. Borrelli, J. L. L. Góngora, R. Kaboteh, et al., “Freely available convolutional neural network-based quantification of pet/ct lesions is associated with survival in patients with lung cancer,” EJNMMI physics 9(1), 1–10 (2022).

5 M. Sadik, J. López-Urdaneta, J. Ulén, et al., “Artificial intelligence increases the agreement among physicians classifying focal skeleton/bone marrow uptake in hodgkin’s lymphoma patients staged with [18f] fdg pet/ct—a retrospective study,” Nuclear Medicine and Molecular Imaging, 1–7 (2022).

6 E. Trägårdh, P. Borrelli, R. Kaboteh, et al., “Recomia—a cloud-based platform for artificial intelligence research in nuclear medicine and radiology,” EJNMMI physics 7(1), 1–12 (2020).

7 M. Antonelli, A. Reinke, S. Bakas, et al., “The medical segmentation decathlon,” (2021).

8 K. W. Clark, B. A. Vendt, K. E. Smith, et al., “The cancer imaging archive (tcia): Maintaining and operating a public information repository.,” J. Digital Imaging 26(6), 1045–1057 (2013).

9 P. Kinahan, M. Muzi, B. Bialecki, et al., “Data from the acrin 6668 trial nsclc-fdg-pet,” (2019).

10 M. Machtay, F. Duan, B. A. Siegel, et al., “Prediction of survival by [18f] fluorodeoxyglucose positron emission tomography in patients with locally advanced non–small-cell lung cancer undergoing definitive chemoradiation therapy: results of the acrin 6668/rtog 0235 trial,” Journal of clinical oncology 31(30), 3823 (2013).

11 H. Roth, L. Lu, A. Seff, et al., “A new 2.5 d representation for lymph node detection in ct,” (2015).

12 H. R. Roth, L. Lu, A. Seff, et al., “A new 2.5 d representation for lymph node detection using random sets of deep convolutional neural network observations,” in International conference on medical image computing and computer-assisted intervention, 520–527, Springer (2014).

13 A. Seff, L. Lu, K. M. Cherry, et al., “2d view aggregation for lymph node detection using a shallow hierarchy of linear classifiers,” in International conference on medical image computing and computer-assisted intervention, 544–552, Springer (2014).

14 N. Heller, N. Sathianathen, A. Kalapara, et al., “C4kc kits challenge kidney tumor segmentation dataset,” (2019).

15 N. Heller, F. Isensee, K. H. Maier-Hein, et al., “The state of the art in kidney and kidney tumor segmentation in contrast-enhanced ct imaging: Results of the kits19 challenge,” Medical Image Analysis 67, 101821 (2021).

16 B. Rister, K. Shivakumar, T. Nobashi, et al., “Ct-org: A dataset of ct volumes with multiple organ segmentations,” (2019).

17 B. Rister, D. Yi, K. Shivakumar, et al., “Ct organ segmentation using gpu data augmentation, unsupervised labels and iou loss,” (2018).

18 P. Bilic, P. F. Christ, E. Vorontsov, et al., “The liver tumor segmentation benchmark (lits),” CoRR abs/1901.04056 (2019).

19 M. Patnana, S. Patel, and A. S. Tsao, “Data from anti-pd-1 immunotherapy lung,” (2019).

20 Ö. Çiçek, A. Abdulkadir, S. S. Lienkamp, et al., “3d u-net: learning dense volumetric segmentation from sparse annotation,” in International conference on medical image computing and computer-assisted intervention, 424–432, Springer (2016).

21 D. P. Kingma and J. Ba, “Adam: A method for stochastic optimization,” arXiv preprint 1412.6980 (2014).

